# Seroprevalence of anti-SARS-CoV-2 IgG antibodies among truck drivers and assistants in Kenya

**DOI:** 10.1101/2021.02.12.21251294

**Authors:** E Wangeci Kagucia, John N Gitonga, Catherine Kalu, Eric Ochomo, Benard Ochieng, Nickline Kuya, Angela Karani, James Nyagwange, Boniface Karia, Daisy Mugo, Henry K Karanja, James Tuju, Agnes Mutiso, Hosea Maroko, Lucy Okubi, Eric Maitha, Hossan Ajuck, Mary Bogita, Richmond Mudindi, David Mukabi, Wycliffe Moracha, David Bulimu, Nelson Andanje, Evans Shiraku, Rosemary Okuku, Monicah Ogutu, Rashid Aman, Mercy Mwangangi, Patrick Amoth, Kadondi Kasera, Wangari Ng’ang’a, Rodgers Mariga, Tobias Munabi, Susan M Ramadhan, Janet Mwikali, Rose Nasike, Cornelius Andera, Roselyne Nechesa, Benson K Kiplagat, Julius Omengo, Simon Oteba, Arthur Mwangi, Dorcas Mkanyi, George Karisa, Judith K Migosi, Patrick Msili, Samson Mwambire, Anthony M Boniface, Amek Nyaguara, Shirine Voller, Mark Otiende, Christian Bottomley, Charles N Agoti, Lynette I Ochola-Oyier, Ifedayo M O Adetifa, Anthony O Etyang, Katherine E Gallagher, Sophie Uyoga, Edwine Barasa, Philip Bejon, Benjamin Tsofa, Ambrose Agweyu, George M Warimwe, J Anthony G Scott

## Abstract

In October 2020, anti-SARS-CoV-2 IgG seroprevalence among truck drivers and their assistants (TDA) in Kenya was 42.3%, higher than among other key populations. TDA transport essential supplies during the COVID-19 pandemic, placing them at increased risk of being infected and of transmitting SARS-CoV-2 infection over a wide geographical area.

## Introduction

Serosurveillance for antibodies to the severe acute respiratory syndrome coronavirus 2 (SARS-CoV-2) can be used to estimate the cumulative incidence of SARS-CoV-2 infection. This is of particular interest among frontline workers, such as truck drivers and their assistants (TDA), who are exempted from coronavirus disease 2019 (COVID-19) restrictions such as curfews and lockdowns.

Shortly after the first locally detected SARS-CoV-2 infection in March 2020, the Government of Kenya instituted containment measures, including travel restrictions[1]. Freight transportation was excluded from travel restrictions as TDA were deemed essential workers [2]. In May 2020 the Kenya Ministry of Health implemented mandatory bimonthly SARS-CoV-2 nucleic acid testing (NAT) for TDA, in line with East African Community (EAC) guidelines[3, 4]. Though NAT data can be used to estimate the cumulative incidence of SARS-CoV-2 infection[5], accurate estimates are dependent on the availability of reliable model parameter values.

There are currently no estimates of SARS-CoV-2 antibody seroprevalence among TDA globally. Due to the nature of their work, TDA interact with a wide variety of social and professional contacts over a large geographical area, putting them at increased risk of SARS-CoV-2 infection. We undertook a serosurvey to estimate the prevalence of SARS-CoV-2 antibodies among TDA in Kenya.

## Methods

### Study design and participants

A cross-sectional serosurvey was conducted at three sites, one in Kilifi County in South-Eastern Kenya, and two in Busia County in Western Kenya (**Supplementary Figure 1**). In Kilifi County, TDA transporting salt domestically and to East African countries (e.g., Uganda, Rwanda and Burundi) were engaged at salt harvesting companies located in Gongoni area, Magarini Sub-County, whilst waiting for salt to be loaded to their trucks. In Busia County, TDA were engaged at One Stop Border Posts (OSBPs) in two border towns, Busia and Malaba as they presented for nasopharyngeal and oropharyngeal (NP/OP) sample collection for NAT as mandated by the Kenyan government. TDA at Busia OSBP and Malaba OSBP typically transport freight between Mombasa on the Indian Ocean Coast, and countries in East Africa (**Supplementary Figure 2**). Busia town is larger and more populous than Malaba[6] yet a higher number of trucks transit through Malaba[7].

TDA were eligible if they were aged ≥18 years without a medical contraindication for blood sample collection. Participation was voluntary and verbal consent was sought prior to blood sample collection. Serosurveillance was conducted as a public health activity requested by the Kenya Ministry of Health and ethical approval for publication of these data was obtained from the Kenya Medical Research Institute Scientific and Ethics Review Unit (KEMRI/SERU/CGMR-C/203/4085).

### Data and sample collection

Sociodemographic and clinical data, including age, sex, nationality, residence, temperature and symptoms of illness within the previous two weeks, were collected by county public health staff (CPHS) and recorded on governmental COVID-19 surveillance forms (**Supplementary Forms 1 and 2**). Venous blood samples were collected in heparinized tubes using standard blood sample collection procedures, labelled with the same anonymous unique identifier as NP/OP samples, then stored in an insulated cool box prior to transportation to the laboratory for processing. NP/OP NAT results, linked using the anonymous unique identifiers, were obtained from testing laboratories. Data with personal identifiers were retained only by CPHS.

### Laboratory testing

Plasma was extracted from blood samples using standard techniques and stored at −80°C prior to antibody testing. Thawed plasma samples were tested for anti-SARS-CoV-2 IgG using a locally-validated whole spike enzyme-linked immunosorbent assay (ELISA) with 93% sensitivity and 99% specificity, as described previously [8, 9]. Spike ELISA positivity was defined as a ratio of sample optical density (OD) over negative control OD > 2. NP/OP NAT results are routinely provided to TDA by CPHS. Serology results were also provided to TDA by CPHS.

### Statistical analysis

At least 700 samples were required to estimate seroprevalence as low as 2% within a 1% margin of error. Age was categorized into 10-year strata. Seroprevalence was estimated as a simple proportion of the number of anti-SARS-CoV-2 IgG positive samples (OD ratio >2) divided by all samples. Associated exact binomial 95% confidence intervals (CI) were calculated for all seroprevalence estimates. Bayesian adjustment for assay sensitivity and specificity was performed as previously described [9]. Associations between seropositivity and sociodemographic/clinical characteristics were tested using chi-squared or Fisher’s exact tests. Bayesian test-performance-adjusted seroprevalence estimates were calculated in R (version 3.6.1) with RStan. All other analyses were performed using Stata (version 15.1).

## Results

The serosurvey was conducted in Magarini between September 30 and October 23, 2020 and in Busia County on October 13-15, 2020. In total, 830 TDA (Busia OSBP n= 365; Magarini n= 101; Malaba OSBP n= 364) provided a blood sample, representing 52.4% of TDA approached (**Supplementary Figure 3**). Of the 830 samples, 91.1% were collected between October 13-15 (**Supplementary Figure 4**). The median age of sampled TDA was 40 years (IQR 34-48 years) and 668 (80.5%) were Kenyan. Only 3 (0.4%) of the TDA were female (**Table 1**). Among Kenyan TDA, reported county of residence included 31 of the country’s 47 counties. The most common counties of residence were Mombasa, Uasin Gishu and Nakuru (**Supplementary Table 1**). Driver vs assistant status was not recorded but, on the basis of truck driver licensure age requirements, at least 53 (6.4%) were assistants.

**Table 1.**
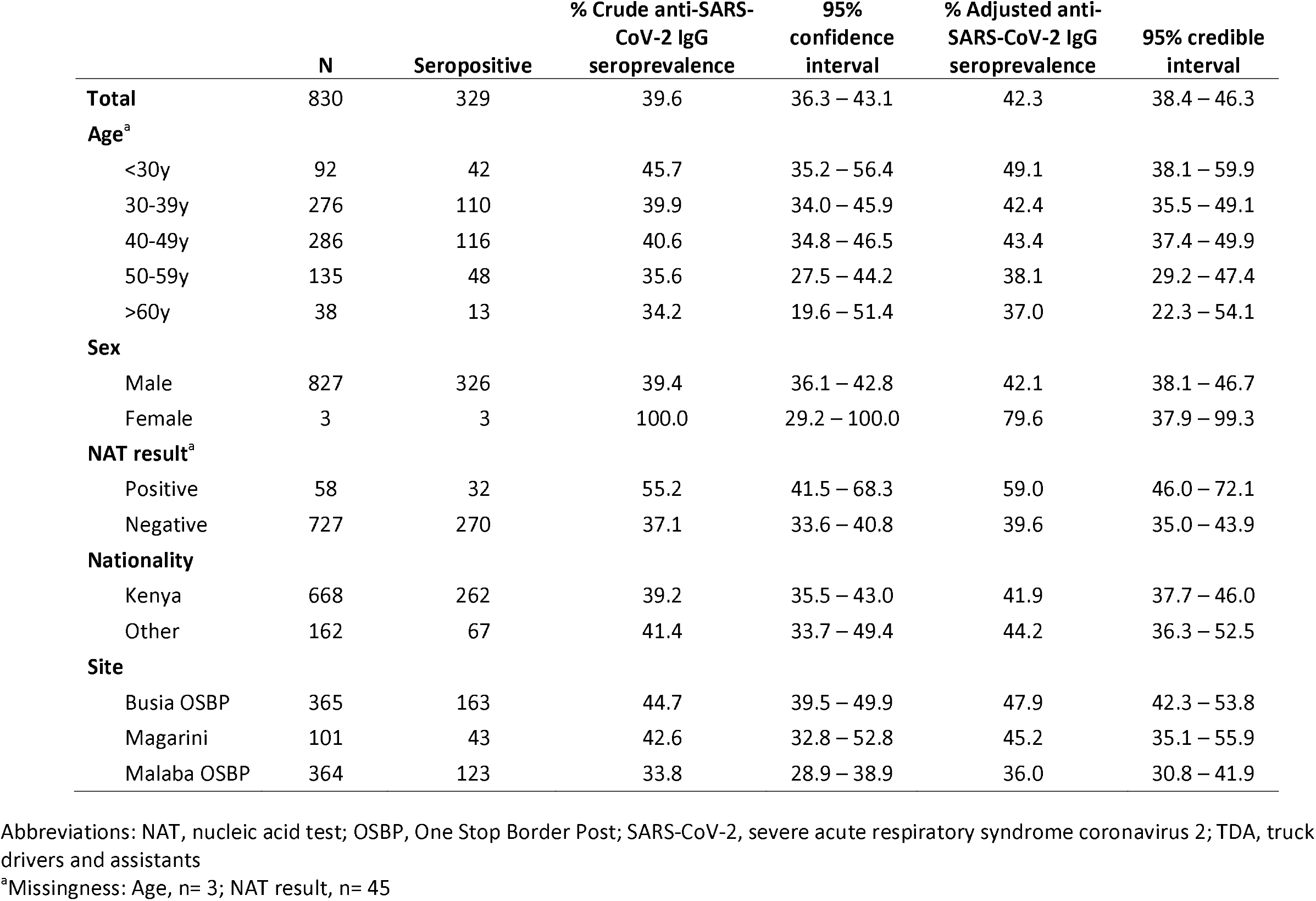
Crude and test-performance adjusted anti-SARS-CoV-2 IgG seroprevalence among TDA in Kenya, September 30 to October 23, 2020

|  | N | Seropositive | % Crude anti-SARS-CoV-2 IgG seroprevalence | 95% confidence interval | % Adjusted anti-SARS-CoV-2 IgG seroprevalence | 95% credible interval |
| --- | --- | --- | --- | --- | --- | --- |
| **Total** | 830 | 329 | 39.6 | 36.3 – 43.1 | 42.3 | 38.4 – 46.3 |
| **Age<sup>a</sup>** |  |  |  |  |  |  |
| <30y | 92 | 42 | 45.7 | 35.2 – 56.4 | 49.1 | 38.1 – 59.9 |
| 30-39y | 276 | 110 | 39.9 | 34.0 – 45.9 | 42.4 | 35.5 – 49.1 |
| 40-49y | 286 | 116 | 40.6 | 34.8 – 46.5 | 43.4 | 37.4 – 49.9 |
| 50-59y | 135 | 48 | 35.6 | 27.5 – 44.2 | 38.1 | 29.2 – 47.4 |
| >60y | 38 | 13 | 34.2 | 19.6 – 51.4 | 37.0 | 22.3 – 54.1 |
| **Sex** |  |  |  |  |  |  |
| Male | 827 | 326 | 39.4 | 36.1 – 42.8 | 42.1 | 38.1 – 46.7 |
| Female | 3 | 3 | 100.0 | 29.2 – 100.0 | 79.6 | 37.9 – 99.3 |
| **NAT result<sup>a</sup>** |  |  |  |  |  |  |
| Positive | 58 | 32 | 55.2 | 41.5 – 68.3 | 59.0 | 46.0 – 72.1 |
| Negative | 727 | 270 | 37.1 | 33.6 – 40.8 | 39.6 | 35.0 – 43.9 |
| **Nationality** |  |  |  |  |  |  |
| Kenya | 668 | 262 | 39.2 | 35.5 – 43.0 | 41.9 | 37.7 – 46.0 |
| Other | 162 | 67 | 41.4 | 33.7 – 49.4 | 44.2 | 36.3 – 52.5 |
| **Site** |  |  |  |  |  |  |
| Busia OSBP | 365 | 163 | 44.7 | 39.5 – 49.9 | 47.9 | 42.3 – 53.8 |
| Magarini | 101 | 43 | 42.6 | 32.8 – 52.8 | 45.2 | 35.1 – 55.9 |
| Malaba OSBP | 364 | 123 | 33.8 | 28.9 – 38.9 | 36.0 | 30.8 – 41.9 |

Abbreviations: NAT, nucleic acid test; OSBP, One Stop Border Post; SARS-CoV-2, severe acute respiratory syndrome coronavirus 2; TDA, truck drivers and assistants
<sup>a</sup>Missingness: Age, n= 3; NAT result, n= 45

Of 785 TDA with available NAT results, 58 (7.4%) were positive for SARS-CoV-2 (**Table 1**) with NAT positivity being significantly higher in Magarini (25 of 93; 26.9%) compared to Busia OSBP (15 of 348; 4.3%) and Malaba OSBP (18 of 344; 5.2%; p <0.001**; Supplementary Table 2**). None of the 830 TDA reported current or previous symptoms of illness. Only one individual had a temperature of 37.5°C or higher (**Supplementary Table 2**) and the NAT result was negative in that one case.

Crude overall anti-SARS-CoV-2 IgG seroprevalence among sampled TDA was 39.6% (95% confidence interval 36.3-43.1). After adjustment for assay sensitivity and specificity it was 42.3% (95% credible interval 38.4-46.3). Seroprevalence was highest in those <30 years of age and lowest among those aged ≥50 years. All three female TDA were seropositive for anti-SARS-CoV-2 IgG. Seroprevalence was 39.6% (35.0-43.9) among TDA negative for SARS-CoV-2 by NAT and 59.0% (46.0-72.1) among those with a positive SARS-CoV-2 NAT result. Seroprevalence among Kenyan TDA was comparable to that of other nationalities. By site, seroprevalence was highest at Busia OSBP and lowest at Malaba OSBP. Crude heterogeneity tests and uncertainty intervals for antibody test-adjusted estimates suggested no significant association between seroprevalence and age and significant differences in seroprevalence by site (**Table 1** and **Supplementary Table 3**). Stratum-specific adjusted seroprevalence estimates were generally higher than crude estimates (**Table 1**).

## Discussion

We estimate that approximately 4 in 10 TDA in Kenya had been infected with SARS-CoV-2 by late October 2020, as the country experienced the initial stages of a second pandemic wave [10]. This study is notable for being the first to report SARS-CoV-2 seroprevalence among TDA.

Anti-SARS-CoV-2 IgG seroprevalence among TDA was more than 4-fold higher compared to average seroprevalence among Kenyan blood donors by September 2020[11] and 2-fold higher compared to average seroprevalence in HCW across five Kenyan health facilities by October 2020 [12]. As such, the findings suggest exposure to SARS-CoV-2 among TDA in Kenya was higher than in the general population and among other frontline workers.

Some studies found seroprevalence of SARS-CoV-2 antibodies among HCW similar to that among TDA in Kenya: 45% in London[13]; 44% in Indiana[14]; and 45% in Ibadan, Nigeria[15]. However, meta-analyses of studies globally estimated SARS-CoV-2 antibody prevalence among HCWs at 7% by July 2020 [16] and 9% by August 2020[17]. Taken together, these data suggest exposure to SARS-CoV-2 among TDA was higher than among HCWs in most settings.

To date, few studies have assessed SARS-CoV-2 antibody prevalence in non-healthcare frontline workers. Seroprevalence was 38% seroprevalence among male migrant supermarket workers in Kuwait in May to June 2020 (during a peak in daily COVID-19 cases)[10, 18]. Elsewhere, however, seroprevalence among non-HCWs has been much lower. SARS-CoV-2 antibody seroprevalence was 1% in Croatia, just prior to easing of nationwide COVID-19 restrictions in late April 2020[19]; 9-11% in France following the initial pandemic wave[20]; 20% in Chicago, Illinois, US during the summer of 2020 following easing of COVID-19 restrictions[21]; and 19-22% in Iran following the initial pandemic wave[22].

This study provides some insights into SARS-CoV-2 epidemiology among TDA. None of the TDA with a positive NAT result reported symptoms within two weeks of testing and no instances of pyrexia were identified among them. This suggests a preponderance of asymptomatic COVID-19 among TDA. Although data on symptoms among TDA may have been subject to reporting bias, 93% of SARS-CoV-2 infections confirmed in Kenya by October 23, 2020 were asymptomatic[23]. The similarity in anti-SARS-CoV-2 antibody seropositivity among Kenyan and non-Kenyan TDA indicates predominantly occupation-related exposure to SARS-CoV-2. That seroprevalence differed by sampling site suggests some heterogeneity in risk of exposure.

Despite sampling at three different sites, we cannot be sure that the sample included are representative of all TDA in Kenya, or even those transiting through these sites. Participation in the serosurvey was voluntary, only half of those approached agreed to be sampled, and sociodemographic data for the target population are not available, limiting our ability to assess recruitment bias. However, data on county of residence for Kenyan TDA indicates representation from most counties. Seroprevalence estimates did not account for clustering by site or by driver-assistant pairs. The use of antibodies to estimate cumulative incidence of SARS-CoV-2 infection assumes universal seroconversion, minimal mortality and antibody persistence. If these assumptions are incorrect – for example, there is evidence suggestive of antibody waning[24–26] – the cumulative incidence of infection would be underestimated. To minimize bias due to assay sensitivity and specificity, adjusted seroprevalence estimates were calculated.

These findings indicate substantial occupational risk for SARS-CoV-2 infection in TDA, despite prevention measures. In turn, infected TDA can transmit SARS-CoV-2 over a large geographical area. Although this study did not assess the contribution of TDA to SARS-CoV-2 transmission, TDA have been previously associated with spatial spread of emerging infections[27–30]. As TDA will continue to play an essential role during the COVID-19 pandemic, heightened promotion of existing COVID-19 prevention measures is needed, as is consideration of prioritization of TDA for COVID-19 vaccines with transmission-blocking potential. Nevertheless, supply chains are essential to sustain daily life and for the COVID-19 response. It remains a challenge how to best balance the essential role of TDA with pandemic control.

## Data Availability

De-identified data may be requested through the KEMRI-Wellcome Trust Research Programme Data Governance Committee

## Abbreviations

COVID-19: Coronavirus disease 2019
CPHS: County public health staff
ELISA: Enzyme-linked immunosorbent assay
HCW: Healthcare workers
NAT: Nucleic acid test/ testing
NP/OP: Nasopharyngeal/ oropharyngeal
OD: Optical density
OSBP: One Stop Border Post
SARS-CoV-2: Severe acute respiratory syndrome coronavirus 2
TDA: Truck drivers and assistants

## Acknowledgments

We thank the truck drivers and assistants for their participation. We thank the Busia Transporters Chairman for support in engaging truck drivers and assistants. We thank the Busia and Kilifi County Health Management Teams (CHMTs), Busia Port Health, Malaba Port Health, and Gongoni area salt companies for their support of this activity, including training support by Busia and Kilifi CHMT members. We thank the Deputy Director, KEMRI Centre for Global Health Research and Deputy Director, KEMRI Centre for Infectious and Parasitic Diseases Control Research for collaborative agreements to support field and laboratory work. We thank the KEMRI-Wellcome Trust Pneumococcal Conjugate Vaccine Impact Study (PCVIS) field staff for supporting sample collection training and data entry. We thank F. Krammer for providing the plasmids used to generate the spike protein used in this work. Development of SARS-CoV-2 reagents was partially supported by the NIAID Centres of Excellence for Influenza Research and Surveillance (CEIRS) contract HHSN272201400008C. This paper has been published with the permission of the Director, Kenya Medical Research Institute.

AA is funded by a DFID/MRC/NIHR/Wellcome Trust Joint Global Health Trials Award (MR/R006083/1), JAGS is funded by a Wellcome Trust Senior Research Fellowship (214320) and the NIHR Health Protection Research Unit in Immunisation, IMOA is funded by the United Kingdom’s Medical Research Council and Department for International Development through an African Research Leader Fellowship (MR/S005293/1) and by the NIHR-MPRU at UCL (grant 2268427 LSHTM). GMW is supported by a fellowship from the Oak Foundation. CNA is funded by the DELTAS Africa Initiative [DEL-15-003], and the Foreign, Commonwealth and Development Office and Wellcome (220985/Z/20/Z). SU is funded by DELTAS Africa Initiative [DEL-15-003], LIO-O is funded by a Wellcome Trust Intermediate Fellowship (107568/Z/15/Z).

*This research was funded in whole or in part by the Wellcome Trust (grants 220991/Z/20/Z and 203077/Z/16/Z)*. ***For the purpose of Open Access, the author has applied a CC-BY public copyright licence to any author accepted manuscript version arising from this submission***.

