## Supplement for "Seroprevalence of anti-SARS-CoV-2 IgG antibodies among truck drivers and assistants in Kenya"

**Supplementary Table 1.** Reported county of residence among Kenyan truckers

| County | Frequency | Percent | County | Frequency | Percent |
|---|---|---|---|---|---|
| MOMBASA | 156 | 23.4 | KIRINYAGA | 4 | 0.6 |
| UASIN GISHU | 76 | 11.4 | LAIKIPIA | 4 | 0.6 |
| NAKURU | 67 | 10.0 | SIAYA | 4 | 0.6 |
| NAIROBI | 60 | 9.0 | KITUI | 3 | 0.5 |
| KIAMBU | 41 | 6.1 | MANDERA | 3 | 0.5 |
| BUSIA | 40 | 6.0 | EMBU | 2 | 0.3 |
| MACHAKOS | 18 | 2.7 | KAJIADO | 2 | 0.3 |
| KWALE | 14 | 2.1 | SAMBURU | 2 | 0.3 |
| BUNGOMA | 13 | 2.0 | GARISSA | 1 | 0.2 |
| KAKAMEGA | 13 | 2.0 | HOMABAY | 1 | 0.2 |
| KISUMU | 13 | 2.0 | KISII | 1 | 0.2 |
| KERICHO | 8 | 1.2 | MIGORI | 1 | 0.2 |
| MAKUENI | 8 | 1.2 | TAITA TAVETA | 1 | 0.2 |
| TRANS NZOIA | 6 | 0.9 | *Uganda* | 1 | 0.2 |
| MURANGA | 5 | 0.8 | *Missing*[a] | 91 | 13.7 |
| NYERI | 5 | 0.8 | Total | 668 | 100 |
| ELGEIYO MARAKWET | 4 | 0.6 | | | |

[a]County of residence not available for 90 of 101 truckers sampled at Magarini due to differences in the data collection form used; County of residence not collected for 1 trucker sampled at Malaba

**Supplementary Table 2**. Distribution of characteristics by site. Tests for heterogeneity conducted using analysis of variance (age) or chi-squared tests (all other variables)

| Characteristic | Busia OSBP N= 365 | | Magarini N= 101 | | Malaba OSBP N= 364 | | p-values |
|---|---|---|---|---|---|---|---|
| **Mean age (SD), range**[a] | 41.0y (9.6) | 20 – 68y | 38.4y (9.3) | 20 - 61y | 42.3y (9.9) | 19 – 78y | 0.002 |
| **Male, %** | 363 | 99.5% | 100 | 99.0% | 364 | 100% | 0.249 |
| **NAT positive, %**[b] | 15 | 4.3% | 25 | 26.9% | 18 | 5.2% | <0.001 |
| **Kenyan, %** | 294 | 80.5% | 73 | 72.3% | 301 | 82.7% | 0.065 |
| **Symptoms, %** | 0 | 0% | 0 | 0% | 0 | 0% | N/A |
| **Temperature ≥$37.5^0$C**[c] | 0 | 0% | 0 | 0% | 1 | 0.3% | 0.547 |

Abbreviations: NAT, nucleic acid testing; OSBP, One Stop Border Post

[a]Missingness: Magarini = 2; Malaba = 1

[b]Missingness: Busia = 17; Magarini = 8; Malaba = 20

[c]Missingness: Busia = 12; Magarini = 17; Malaba = 1

**Supplementary Table 3**. Stratum-specific tests for heterogeneity using crude anti-SARS-CoV-2 IgG seroprevalence

| | N | Seropositive | % Crude anti-SARS-CoV-2 IgG seroprevalence | 95% CI | p-value |
|---|---|---|---|---|---|
| **Age**[a] | | | | | |
| <30y | 92 | 42 | 45.7 | 35.2 – 56.4 | 0.144[a] |
| 30-39y | 276 | 110 | 39.9 | 34.0 – 45.9 | |
| 40-49y | 286 | 116 | 40.6 | 34.8 – 46.5 | |
| 50-59y | 135 | 48 | 35.6 | 27.5 – 44.2 | |
| >60y | 38 | 13 | 34.2 | 19.6 – 51.4 | |
| **Sex** | | | | | |
| Male | 827 | 326 | 39.4 | 36.1 – 42.8 | 0.062[b] |
| Female | 3 | 3 | 100.0 | 29.2 – 100.0 | |
| **NAT result** | | | | | |
| Positive | 58 | 32 | 55.2 | 41.5 – 68.3 | 0.007 |
| Negative | 727 | 270 | 37.1 | 33.6 – 40.8 | |
| **Nationality** | | | | | |
| Kenya | 668 | 262 | 39.2 | 35.5 – 43.0 | 0.618 |
| Other | 162 | 67 | 41.4 | 33.7 – 49.4 | |
| **Site** | | | | | |
| Busia OSBP | 365 | 163 | 44.7 | 39.5 – 49.9 | 0.009 |
| Magarini | 101 | 43 | 42.6 | 32.8 – 52.8 | |
| Malaba OSBP | 364 | 123 | 33.8 | 28.9 – 38.9 | |

Abbreviations: NAT, nucleic acid test; OSBP, One Stop Border Post; SARS-CoV-2, severe acute respiratory syndrome coronavirus 2; TDA, truck drivers and assistants

[a]Chi-square test for trend performed

[b]Fisher's exact test performed

**Supplementary Figure 1.** Map of Kenya showing the serosurvey sites

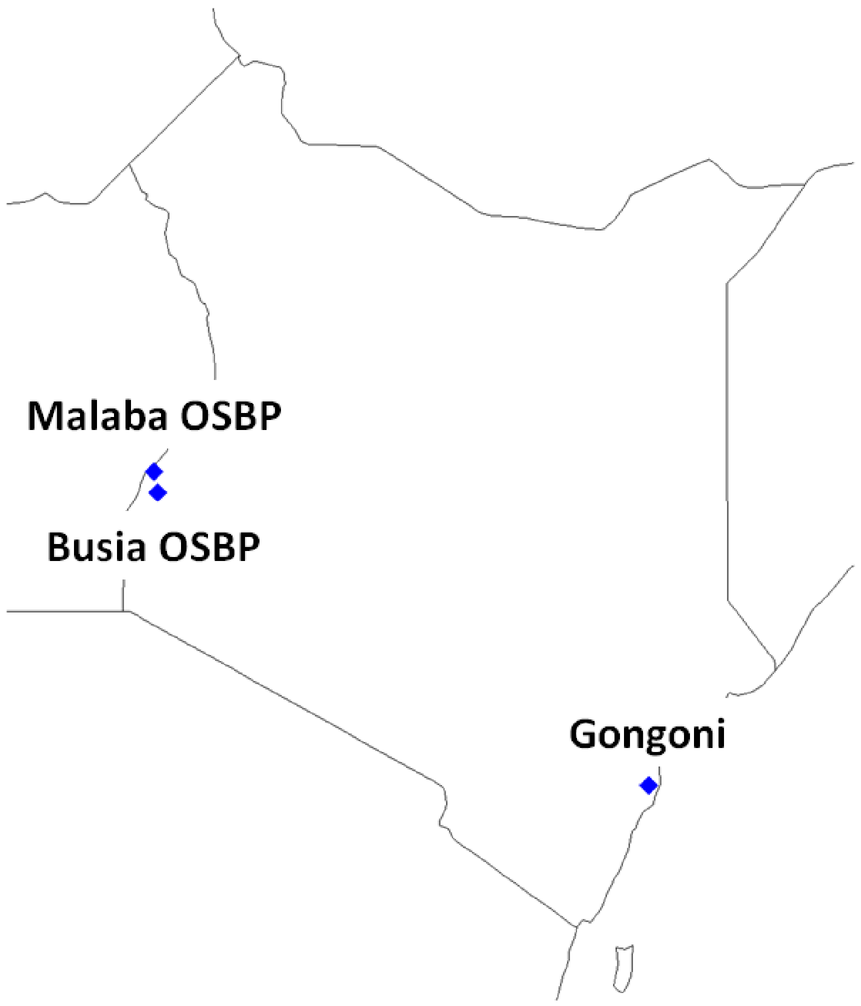

**Supplementary Figure 2.** Map showing the Northern Corridor road network which is used to transport freight from Mombasa, Kenya to other East African countries through Busia OSBP and Malaba OBSP, among other ports. Map from the Northern Corridor Transit and Transport Coordination Authority.[1]

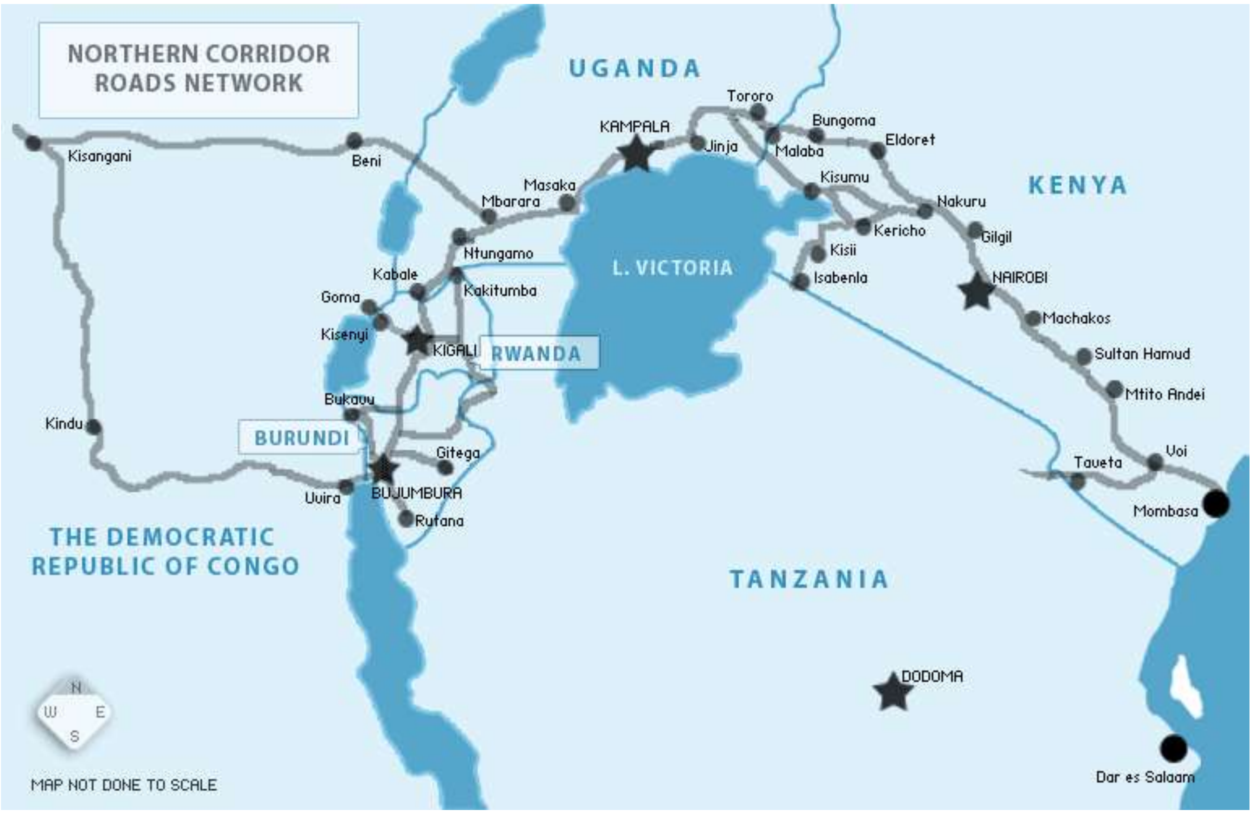

**Supplementary Figure 3.** Flow of TDA included in the SARS-CoV-2 serosurvey

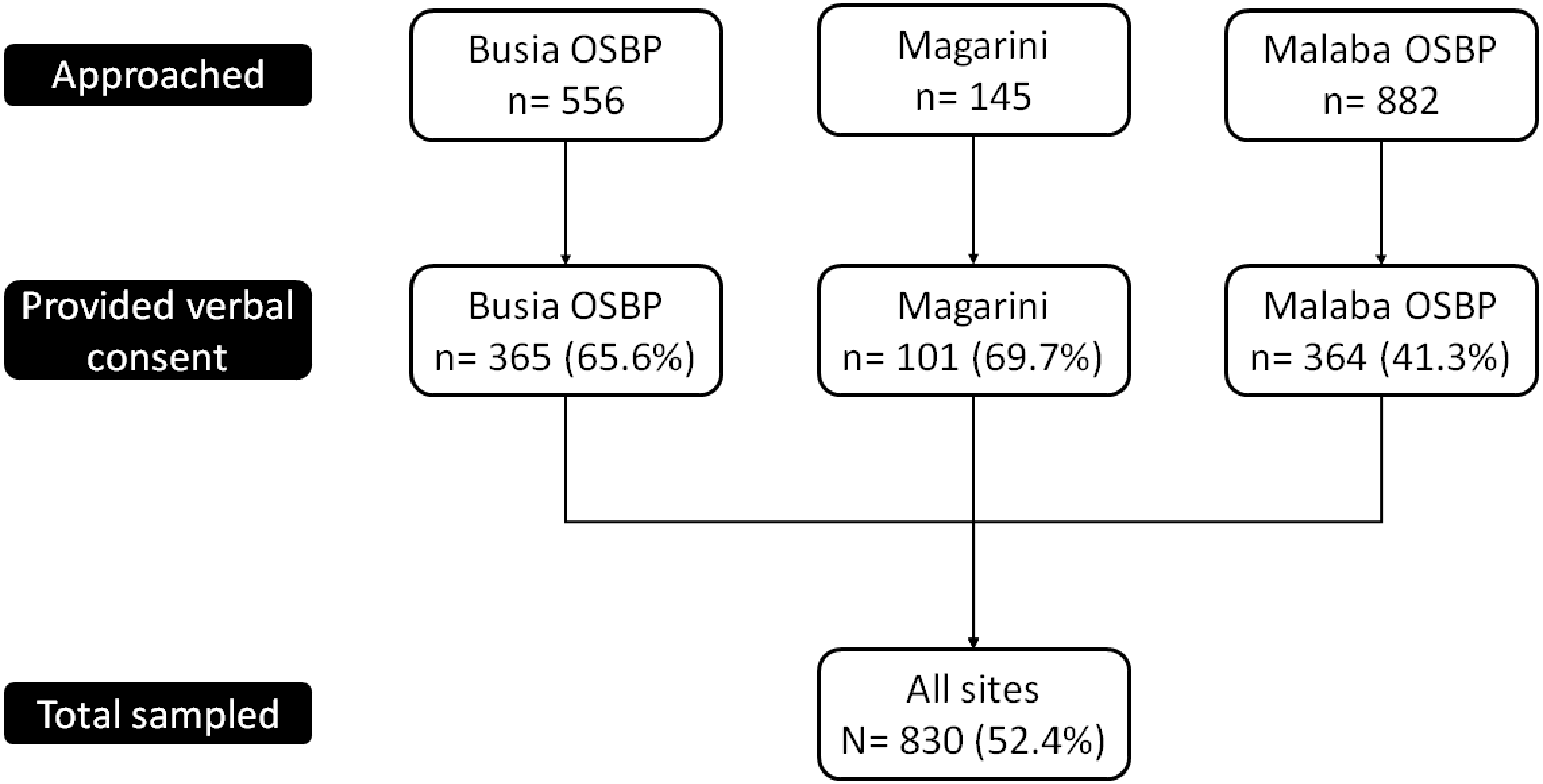

**Supplementary Figure 4.** Sampling by date and site

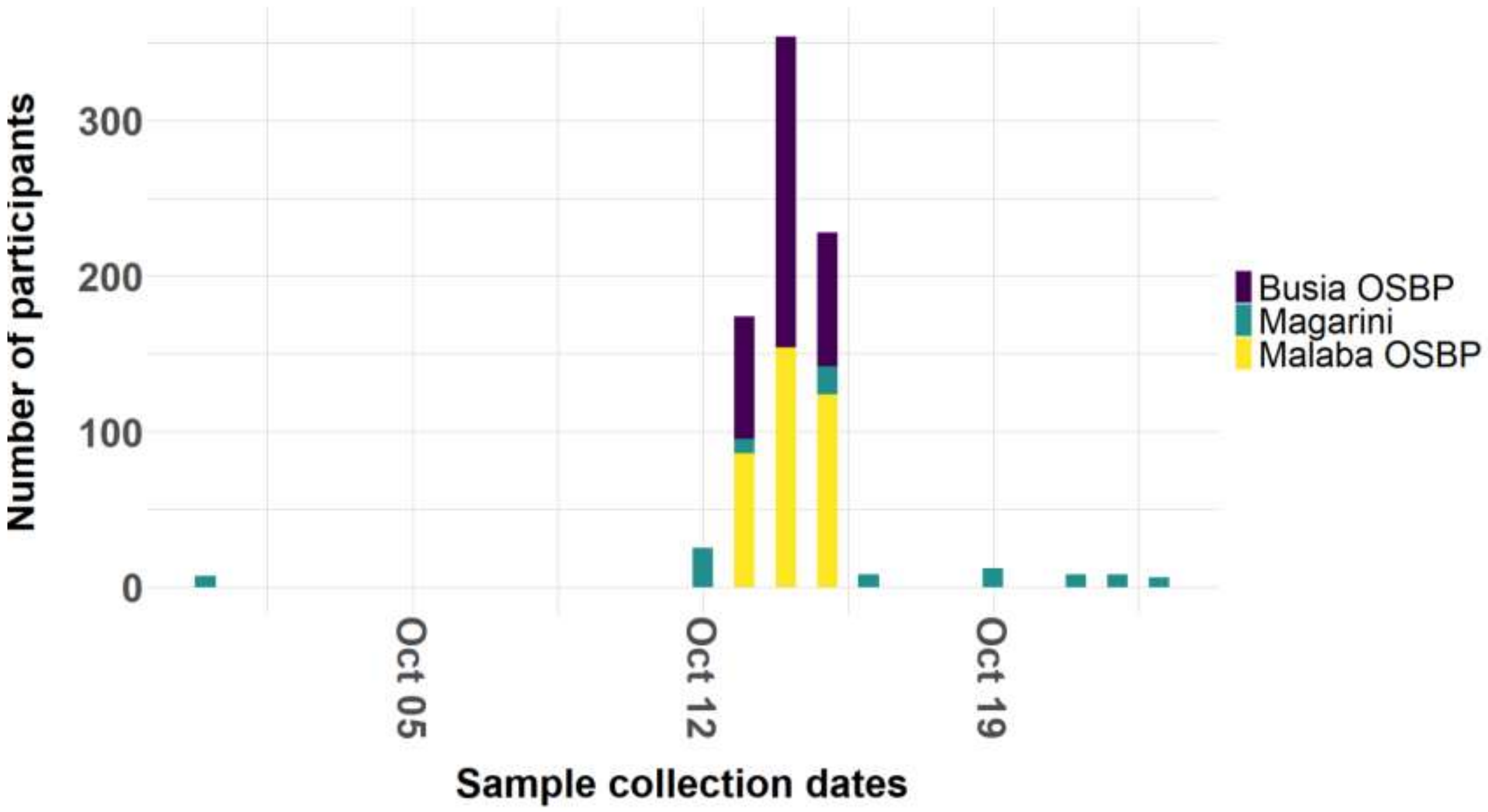

**Supplementary Form 1**

V4

**Test Justification**
Baseline ☐
Follow-up ☐

**MINISTRY OF HEALTH**
**Division of Disease Surveillance and Surveillance**

### Case investigation form for 2019 Novel Coronavirus (COVID-19)

Date of reporting to national level: D[__][__]/M[__][__]/Y[__][__][__][__]

Why was the person tested for COVID-19 or investigation being conducted?
- ☐ Contact with confirmed case
- ☐ Surveillance
- ☐ Repatriation
- ☐ Presented at health facility
- ☐ Point of entry detection
- ☐ Other

Date of investigation:
D[__][__]/M[__][__]/Y[__][__][__][__]

**Section 1: Patient information**

1.1 Unique Case Identifier (used in county): ____________________________

1.2 Full name: ____________________________________________________

1.3 Nationality: ___________ Contact Phone Number: _______________

1.4 Citizenship: ______________ Email address: _______________________

1.5 Age: [__][__] in years

1.6 Sex: ☐ Male ☐ Female

1.7 Place where the case was diagnosed: ☐ Health Facility ☐ Household

County:____________ Sub county: __________________Ward:__________

1.7.1 If health facility, name of health facility: ______________

1.7.2 Patient usual place of residence (village/estate): _______________

**Section 2: Clinical information**

**Patient clinical course**

2.1 Date of onset of symptoms: D [__] [__]/M [__] [__]/Y [__] [__] [__] [__]

☐ Asymptomatic ☐ Unknown

2.2 Admission to hospital: ☐ No ☐ Yes ☐ Unknown

2.2.1 If yes, first date of admission to hospital:

D[__][__]/M[__][__]/Y[__][__][__][__]

2.2.2 Name of hospital: ___________________

2.2.3 Patient taken to isolation ☐ No ☐ Yes ☐ Unknown

1

If yes, date of isolation: D [__][__]/M[__][__]/Y[__][__][__][__]

2.2.4 Patient admitted to ICU ▫ No ▫ Yes ▫ Unknown
2.2.5 Was the patient ventilated: ▫ No ▫ Yes ▫ Unknown
2.3 Health status at time of reporting:
▫ Stable ▫ Severely ill ▫ Dead ▫ Unknown
2.3.1 Outcome ▫ Still in hospital ▫ Discharged ▫ Death
2.3.2 Date of outcome (discharged, death) if applicable:
D [__][__]/M [__][__]/Y[__][__][__][__]

**2.4 Patient symptoms** (check all reported symptoms):

| | | |
|---|---|---|
| ▫ History of fever / chills | ▫ Shortness of breath | ▫ Pain (check all that apply) |
| ▫ General weakness | ▫ Diarrhea | *( ) Muscular ( ) Chest* |
| ▫ Cough | ▫ Nausea/vomiting | *( ) Abdominal ( ) Joint* |
| ▫ Sore throat | ▫ Headache | |
| ▫ Runny nose | ▫ Irritability/Confusion | |

▫ Other, specify ______________________________

Have the symptoms resolved? Yes ☐ No ☐ Unknown ☐

If Yes, Date of symptom resolution ____________________ Unknown ☐

**Patient signs**:
2.5 Temperature ________ ⁰C
2.6 Check all observed signs:

| | | |
|---|---|---|
| ▫ Pharyngeal exudate | ▫ Coma | ▫ Abnormal lung X-Ray findings |
| ▫ Conjunctival injection | ▫ Dyspnea / tachypnea | |
| ▫ Seizure | ▫ Abnormal lung auscultation | |

▫ Other, specify: ______________________________

**2.7 Underlying conditions and comorbidity** (check all that apply):

| | |
|---|---|
| ▫ Pregnancy (trimester:_________) | ▫ Post-partum (< 6 weeks) |
| ▫ Cardiovascular disease, including hypertension | ▫ Immunodeficiency, including HIV |
| ▫ Diabetes | ▫ Renal disease |
| ▫ Liver disease | ▫ Chronic lung disease |
| ▫ Chronic neurological or neuromuscular disease | ▫ Malignancy |

▫ Smoking (current or former smoker)
▫ Other, specify: ______________________________

### Section 3: Exposure and travel information in the 14 days prior to symptom onset (prior to reporting if asymptomatic)

**3.1 Occupation**: (tick any that apply)
▫ Student ▫ Health care worker, Cadre______________________
▫ Working with animals ▫ Health laboratory worker ▫ Other, specify: _______
3.2 Has the patient **travelled** in the 14 days prior to symptom onset? ▫ No ▫ Yes ▫ Unknown
If yes, please specify the places the patient travelled:

| | Country | City | Date |
|---|---|---|---|
| 1 | ______ | ______ | ______ |
| 2 | ______ | ______ | ______ |

3.3 Has the patient **visited any health care facility**(s) in the 14 days prior to symptom onset?

□No □ Yes □ Unknown

3.4 Has the patient had **close contact**[1] with a person with acute respiratory infection in the 14 days prior to symptom onset?

□ No □ Yes □ Unknown

If yes, contact setting (check all that apply):

□ Health care setting □ Family setting □ Workplace □ Unknown

□ Other, specify: ______________________

3.5 Has the patient **had contact with a probable or confirmed case** in the 14 days prior to symptom onset?

□ No □ Yes □ Unknown

If yes, please list unique case identifiers of all probable or confirmed cases:

Case 1 identifier.____________ Case 2 identifier.______________ Case 3 identifier. ______________

If yes, contact setting (check all that apply):

□ Health care setting □ Family setting □ Workplace □ Unknown □Other, specify: ______________

If yes, location/city/country for exposure: ______________________

### Section 4: Laboratory Information

**Specimen collection** *(To be completed by the health facility)*

4.1 Was specimen collected? ☐ 1=Yes 2=No

If no, why?

.......................................................................................................................................

4.2 Date(s) of specimen collection: D[__][__]/M[__][__]/Y[__][__][__][__]

4.3 Specimen type: ☐ NP Swab ☐ OP Swab ☐ Serum ☐ Sputum ☐ Tracheal Aspirate

Other (specify): ______________________

4.4 Date specimen send to the lab: D[__] [__]/M[__][__]/Y[__][__][__][__]

***(To be completed by the confirming lab)***

4.5 Date specimen received in the lab: D[__][__]/M[__][__]/Y[__][__][__][__]

Time[__][__]:[__][__]

4.6 Name of confirming lab: ______________________________

4.7 Please specify which assay was used:____________________

4.8 Preliminary lab results: ______________________________

4.9 Has sequencing been done? □ Yes □ No □ Unknown

4.10 Date of laboratory confirmation: D[__][__]/M[__][__]/Y[__][__][__][__]

[1]Close contact' is defined as: 1. Health care associated exposure, including providing direct care for COVID-19 patients, working with health care workers infected with novel coronavirus, visiting patients or staying in the same close environment of a COVID-19 patient. 2. Working together in close proximity or sharing the same classroom environment with a with COVID-19 patient. 3. Traveling together with COVID-19 patient in any kind of conveyance. 4. Living in the same household as a COVID-19 patient

### Supplementary Form 2

**MINISTRY OF HEALTH**

**COVID – 19 Laboratory Request Form**

**Version 2.0**

Site Name/Facility________________________________Contact Person_______________ Phone# __________________ Date of Dispatch ___________________

Email Address ____________________________________ County ______________________ Sub-County __________________________________

Date Sample Received ___________________________________________ Shipment Temp. Max ___________ Min ______________ Ambient _________

Collected By: ____________________________________________________Date of Sample Collection ________________________________

| # | Case ID/ | Type of Test Initial/ Repeat | Sample No | Full Name | ID PP No | Temp | Age | Sex M/F | Phone No. | Occupation | Nationality | County Residence | Sub-County Of Residence | Village/ Estate | Contact With case | Date Symptom Onset | Symptoms (Cough, Fever, DIB etc) | Type of Sample( OP/NP/ Blood) |
|---|---|---|---|---|---|---|---|---|---|---|---|---|---|---|---|---|---|---|
| 1 | | | | | | | | | | | | | | | | | | |
| 2 | | | | | | | | | | | | | | | | | | |
| 3 | | | | | | | | | | | | | | | | | | |
| 4 | | | | | | | | | | | | | | | | | | |
| 5 | | | | | | | | | | | | | | | | | | |
| 6 | | | | | | | | | | | | | | | | | | |
| 7 | | | | | | | | | | | | | | | | | | |
